# Resection of highly functionally connected glioma regions predicts long-term cognitive preservation

**DOI:** 10.64898/2026.09.28.26363885

**Authors:** Marike R van Lingen, Amit Jaiswal, Srikantan S Nagajaran, Velmurugan Jayabal, Martin Klein, Niels Verburg, Philip de Witt Hamer, Arjan Hillebrand, Linda Douw

## Abstract

**Background:** Glioma surgery requires balancing maximal tumor resection against preservation of neurological and cognitive function. Intraoperative direct electrical stimulation guides this balance but is invasive and not always feasible. Preoperative magnetoencephalography (MEG) can identify tumor regions with high functional connectivity (HFC) to the rest of the brain. These HFC areas contain more malignant glioma cells and relate to poorer short-term outcome when resected. We aimed to determine whether HFC voxel resection relates to postoperative neurological and cognitive outcomes.

**Methods:** In this preregistered analysis, 54 adults with diffuse glioma underwent resting-state MEG before resection. Imaginary coherence identified voxels with significantly higher connectivity against their contralateral controls, and these HFC voxels within the resection cavity were counted. Neurological status was assessed at short-term (∼1 week) and long-term (∼1 year) follow-up, and neuropsychological assessment took place at baseline and long-term follow-up. Logistic regressions related resected HFC voxels to decline at each timepoint, adjusting for Karnofsky performance status, sex and resectability index.

**Results:** Resected HFC voxels were not associated with neurological decline at short-term (48% declined; odds ratio [OR] 0.98, *P* = .50) or long-term follow-up (27% declined; OR 0.99). For cognition, more resected HFC voxels were associated with less long-term decline (48% declined; OR 0.85, *P* = .02); a binarized analysis was consistent (OR 0.076, *P* = .01), indicating that tumor volume did not drive this result.

**Conclusions:** Resecting more HFC voxels associates with preserved long-term cognition, without apparent neurological cost. Non-invasive MEG connectivity mapping may help optimize the onco-functional balance in glioma surgery.

**Key points:**

- Resecting more highly functionally connected tumor voxels relates to preserved long-term cognition in glioma.
- Connectivity did not predict postsurgical neurological outcome.
- Preoperative MEG connectivity mapping may help inform the onco-functional balance.

**Importance of the Study:** Achieving the optimal onco-functional balance in glioma surgery means resecting as much tumor as possible, while preserving neurological and cognitive function over the long term. Intraoperative direct electrical stimulation helps guide this balance but is invasive, time-consuming, and not always feasible. This study extends preoperative, non-invasive MEG functional connectivity mapping to neurological and cognitive outcomes up to one year after surgery, using a different MEG system, cohort, and center than used in prior work. We show that resecting more high functionally connected voxels associates with preserved long-term cognition, at no neurological cost. Preoperative MEG connectivity could therefore complement stimulation mapping to guide safer, more effective resections, particularly in IDH-mutant glioma, whose longer survival makes long-term cognitive preservation especially important.

## Introduction

Surgical resection of glioma consists of finding a delicate balance between maximally resecting the tumor for survival benefits and preserving postoperative neurological and cognitive functioning. Intraoperative mapping techniques, such as direct electrical stimulation (DES) combined with e.g., language or sensory mapping, are routinely performed during surgery, and seem to have improved this onco-functional balance.^1^ However, most studies looking into functional outcomes have focused on short-term neurological and cognitive functioning. Moreover, DES is not always successful, can be time-consuming and may be stressful for patients and healthcare professionals. Preoperative, non-invasive techniques that allow for longer-term outcome prediction, therefore, remain of paramount interest.

Connectivity mapping based on resting-state magnetoencephalography (MEG) may be such an approach. A considerable body of work from a large clinical MEG program has shown that glioma may present with particular patterns of functional connectivity (FC) between the tumor area and the rest of the brain,^2^ with functional connectivity referring to the statistical dependencies between time series of brain activity measured at different locations. This research shows that levels of FC measured preoperatively correlate well with DES functional mapping.^3–5^ Particularly high FC (HFC) tumor voxels are of interest, as they seem to be more malignant from an oncological perspective, at least in glioblastoma.^2^ They also associate with poorer short-term neurological and language outcomes when resected,^6,7^ particularly in DES-negative cases.^7^ However, it remains unknown whether these findings translate to other MEG systems and centers. Moreover, the relation of preoperative HFC patterns to neurological and cognitive outcomes in the longer term, i.e., more than 6 months after resection, remain unexplored. This is all the more of interest, as it becomes clear that more intricate relationships between the tumor and the rest of the brain are prognostically detrimental.^2^

We therefore aimed to partially replicate previous studies on the relationship between preoperative HFC and neurological outcome, and to expand the postoperative outcome measures to neurological and cognitive status up to one year after surgery, in light of their relevance to patients’ quality of life. We hypothesized that resection of more HFC voxels would be related to greater postoperative neurological and cognitive decline in glioma patients, as compared to fewer HFC voxels resected.

## Methods

### Participants

Patients participated in an ongoing prospective observational study at the outpatient clinic of Amsterdam UMC between 2010 and 2021. Study visits took place before resective surgery or any other oncological treatment (baseline, BL) and at approximately 1 year after surgery (long-term follow-up, LFU), with an additional short-term follow-up (SFU) for neurological status taking place clinically at discharge approximately 1 week after surgery. Patients who showed tumor progression before LFU were excluded from the long-term outcome analyses, but remained included in the SFU analyses.

Inclusion criteria were: (1) adult (>17 years old), (2) isocitrate dehydrogenase (IDH)-wildtype glioblastoma, IDH-mutant non-codeleted astrocytoma, or IDH-mutant 1p/19q-codeleted oligodendroglioma based on the 2021 WHO classification,^8^ (3) resting-state MEG performed at BL, (4) neuropsychological assessment (NPA) available at both BL and LFU. Exclusion criteria were: (1) psychiatric disease, (2) comorbidities of the central nervous system (e.g., cerebrovascular accidents, Alzheimer’s or Parkinson’s disease), (3) insufficient mastery of the Dutch language, and (4) inability to communicate adequately. Additional exclusion criteria for the current analyses were (1) bilateral tumor location, and (2) biopsy instead of resective surgery. All participants gave written informed consent before inclusion, and the study was approved by the VUmc Medical Ethical Committee and conducted according to the Declaration of Helsinki.

Part of this data has been published previously (see Supplementary Table 1), using different research questions and analysis methods. The current analysis was preregistered prior to performing any data analysis (https://osf.io/nqetx/).

### Neurological outcome

Neurological examination and scoring of the Karnofsky Performance Status (KPS)^9^ were performed by treating physicians at all three timepoints (BL, SFU, and LFU) as part of routine clinical care. Neurological examination findings were retrospectively extracted from patients’ medical records by our study team, and scored using a standardized scale ranging between 0 and 42, assessing amongst others visual fields, motor and sensory functioning of the arms and legs, language functioning, etc.^10^ Neurological decline between BL and SFU, as well as BL and LFU, was defined as Δ score > 0.

### Cognitive outcome

Neuropsychological assessments were conducted at BL and LFU. The cognitive test battery has been described previously,^11^ and comprises the domains of verbal learning and recall using the Rey Auditory Verbal Learning Test; selective attention, mental concentration, memory, and information processing using the Memory Comparison Test; lexical access and updating aspects of executive functioning using the Categoric Word Fluency test; attention and inhibition using the Stroop-Color Word Test; attention, working memory, and set shifting using the Concept Shifting Test; and information processing speed using the Letter Digit Substitution Test. Each subtest score was converted to a Z-score using validated normative data, thereby adjusting for age, sex, and education.^12,13^ At BL, cognitive impairment was defined as Z-scores < -1.5 on two or more subtests, following the International Cognition and Cancer Task Force recommendation.^14^

Then, changes in Z-scores were calculated for all 13 subtests (LFU - BL). Cognitive decline was defined as ΔZ ≤ -1 on at least 4 subtests following previous work.^11^ Patients with more than 6 cognitive subscores missing on either timepoint were excluded from the cognitive outcome analyses.

### Tumor and cavity segmentation

Preoperative and postoperative MRIs were collected as part of clinical care; T1-weighted images with and without contrast injection, fluid-attenuated inversion recovery (FLAIR) images, and T2-weighted images were included. Tumor masks were segmented using an automated deep-learning-based toolbox, picture-nnunet, that was trained over >1200 manual tumor segmentations of low- and high-grade gliomas(^15^; gitlab.com/picture-production/picture-nnunet-package). The toolbox segmented three tumor classes: (1) edema / non-enhancing tumor, (2) necrotic core / non-enhancing solid tumor and (3) enhancing tumor. We binarized these combined subclasses into one preoperative tumor mask. A tumor occurrence map for all patients was created by coregistering all tumor masks to the Montreal Neurological Institute (MNI) template brain.

The resection cavity was segmented using a postoperative deep-learning algorithm.^16^ Both the tumor and resection cavity masks were visually inspected and manually corrected [by MvL] using 3D Slicer (version 5.2.2) when necessary. The resection cavity mask was mapped to the preoperative T1-weighted MRIs in order to later be overlaid with the FC map. This was done using the linear registration tool FLIRT (FMRIB’s Linear Image Registration Tool, version 6.0) or non-linear registration (FNIRT) in case of skull deformation due to resective surgery. If FNIRT still did not suffice, the next available postoperative T1 scans (max. 9 months after surgery) were used to obtain the resection cavity and register to the preoperative T1 scan. Finally, the tumor and resection cavity masks were merged into one single binary mask, followed by a 1–2 mm dilation to include voxels at the edges. These *merged* masks were used instead of the tumor masks alone to examine FC for all potentially resected voxels, as the resected volume did not always fully overlap with the tumor volume.

### Magnetoencephalography

MEG was recorded in the supine position during a 5-minute no-task eyes-closed resting state at sampling frequency 1250 Hz with a 306-channel VectorView^TM^ system (Elekta Neuromag Oy, Helsinki, Finland, 102 magnetometers, 204 gradiometers) housed in a magnetically shielded room (VacuumSchmelze GmBh, Hanau, Germany). Anti-aliasing (410 Hz) and high-pass filters (0.1 Hz) were applied online. Patients’ head positions were recorded with four or five head position coils, which were digitized together with scalp shape and three fiducial landmarks before the recording, using a 3D digitizer (Fastrak; Polhemus, Colchester, VT, USA). Recordings were visually checked for bad channels or artifacts, after which the spatiotemporal signal space separation method (tSSS;^17^) with a window length of 10 seconds and correlation limit of 0.9 was applied using MaxFilter^TM^ (version 2.0; Megin Oy, Espoo, Finland).^18^

Experimental MEG2map software (version 1.10), developed as a research tool by MEGIN and not commercially available, was used for further preprocessing of the MEG data, following the general analysis pipeline as developed in UCSF^2,6^. MEG2map is Python-based software employing preprocessing, forward modeling, beamforming, and functional connectivity pipelines using customized Python routines employing MNE-Python^18^ (version 1.10.2), Scikit-learn (version 1.5.1), Nibabel (5.3.3), Trimesh (version 4.4.3), and other Python dependencies. It also integrated the robust selection of contralateral control region, statistical testing, data visualization approaches, and detailed reports of intermediate and final results. The software can be used both in graphical and command-line mode on a Linux system.

The tSSS-processed data were bandpass filtered using a fourth-order Butterworth zero-phase non-causal IIR filter for a 1–20 Hz frequency range. The filtered data were downsampled to 600 Hz and an approximately 1-minute-long artifact-free segment was selected for further analysis. MEG data and noise covariance matrices were computed for defining spatial filters. The covariance was computed based on this selected data segment, while the noise covariance was computed using the closest (<15 days apart) recorded empty-room MEG data after applying the same preprocessing steps.

For source localization, preoperative T1 scans were segmented using FreeSurfer (version 7.4.1).^19^ The inner skull, outer skull, and scalp surfaces were computed from the segmented MRI using *mri_watershed* and *mkheadsurf* routines. An MEG-MRI coregistration transform was computed by registering the scalp digitization points from MEG with the MRI scalp surface using the ICP (iterative closest point)-based approach followed by manual adjustments. The potential defects in the inner-skull surface due to the tumor were corrected using a customized Python pipeline and the corrected inner-skull surface (icosahedron, subdivision 5) was used for defining a single-layer volume conductor model using the BEM method as implemented in MNE-Python.^18^ A volumetric source model was defined by placing an equispaced rectangular grid of 8 mm resolution over the inner-skull surface, resulting in approximately 3500 dipolar sources (voxels) inside the brain. The forward solution (leadfield) was computed for each dipolar source in three orientations, using the BEM and source models, the MEG-MRI transform, and the locations and orientations of the MEG sensors.

A scalar-type linear-constrained minimum variance (LCMV) spatial filter (beamformer) was computed for each dipolar source using the precomputed data covariance, noise covariance, and the forward solution in the optimized orientation, which was the one producing the maximum response.^21^ The source time series for each dipolar source was reconstructed from the 1-minute-long segment of data by projecting it through the spatial filters. Subsequently, the power spectrum density (PSD) for the source time series was calculated using the short-time fourier transform (STFT) with a 1-second Hanning window. For each patient, the peak alpha frequency within the 7-14 Hz frequency range was identified, and a range of 2.5-3.0 Hz around these individual alpha peaks was manually selected for the alpha-band connectivity computation.

### Functional connectivity mapping

Similar to previous work,^2,6^ the imaginary coherence^22^ (IC) was used to estimate FC for the selected frequency range around the peak alpha frequency (frequency resolution = 1.17 Hz). First, the dipolar sources inside the combined tumor/cavity mask, hereafter referred to as *tumor voxels*, were identified (see Figure 1). Additionally, a set of dipolar sources contralateral to the tumor voxels, hereafter referred to as *control voxels*, were identified to serve as control voxels. Finally, a subsampled set of every fourth voxel from the entire set of voxels was selected, referred to as *reference voxels.* The IC was computed between each of the tumor voxels and the reference voxels as well as between each of the control and the reference voxels, resulting in 10000 – 100000 values. The IC values were transformed to their absolute values and subsequently averaged over the frequency bins,^2,6,22^ followed by a Z-transformation. Thus, each tumor and reference voxel was associated with a distribution of Z-transformed connectivity values with the reference voxels.

**Figure 1.**
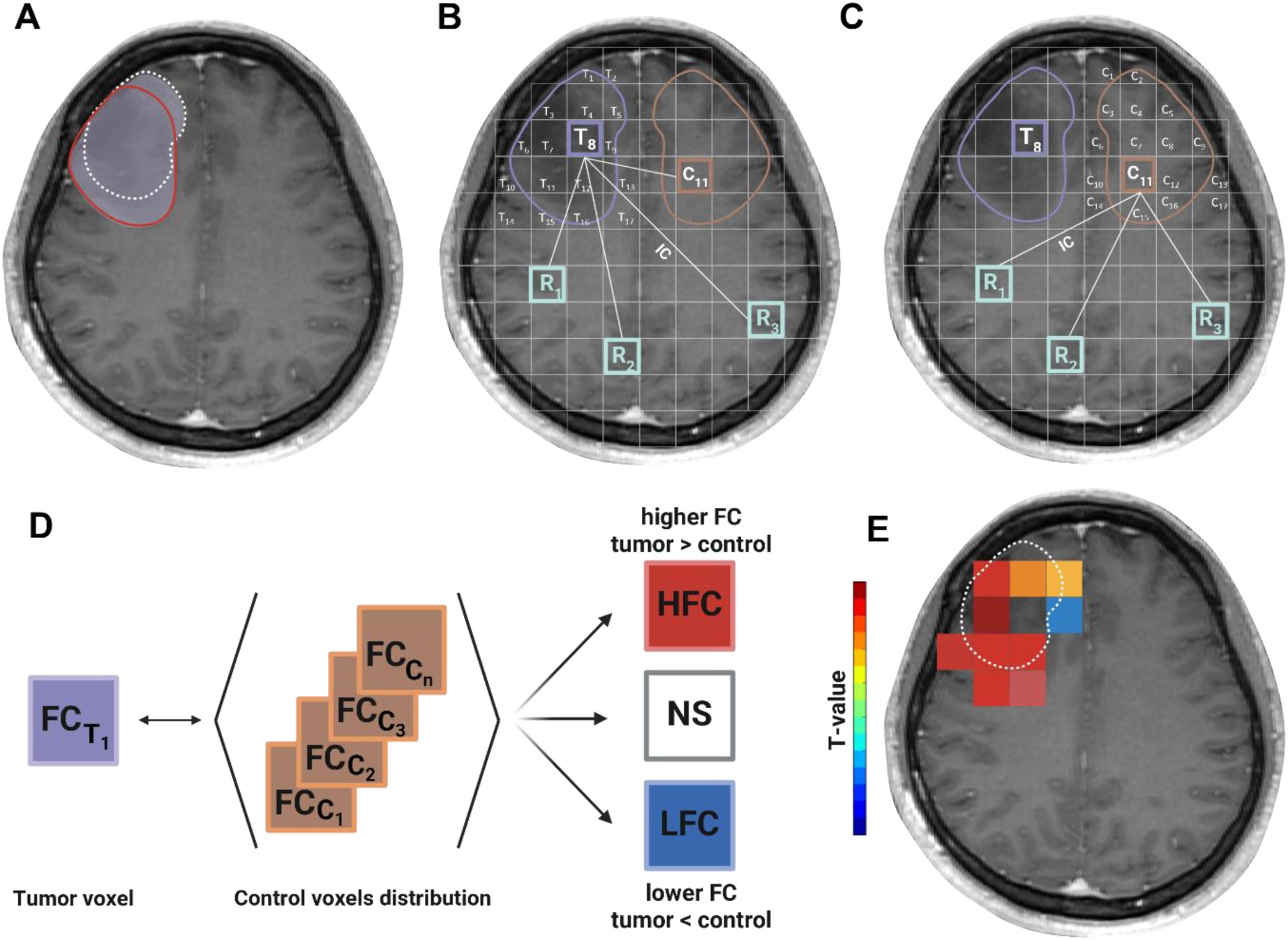
Schematic overview of steps leading to MEG-based functional connectivity (FC) map. **A)** The tumor mask (outlined in red) and the resection cavity mask (white dotted line) were merged into one mask (filled light purple) shown on the preoperative 3D-T1 MRI scan. **B)** FC was calculated using imaginary coherence between each tumor voxel (8 mm^3^) and the reference voxels (each of the fourth voxels from the source space) including contralateral control voxels (C1) and reference voxels in the rest of the brain (R1/R2/R3) . **C)** FC between all control voxels and all reference voxels. **D)** Non-parametric T-test to test if a given tumor voxel has a significantly higher (HFC) or lower (LFC) imaginary coherence distribution compared to the imaginary coherence distribution of all the control voxels. Voxels with non-significantly (NS) different imaginary coherence were excluded from the FC map. **E)** Significant HFC and LFC are mapped onto the preoperative scan. The resected HFC/LFC voxels were identified by overlapping the FC map with the cavity mask. The total number of HFC/LFC/NS voxels is the union of all voxels within the merged mask. The color bar indicates the T-value of each individual voxel.

Non-parametric t-tests were performed to compare the IC distribution of each tumor voxel against the distribution of control voxels, to test which tumor voxels had higher or lower FC than their homologue control voxel (Figure 1). The resulting t-statistics map and the merged mask boundary were overlaid on the preoperative T1 scan. For the final outcome measure, we quantified the number of significantly (*P* < .05) highly functionally connected (HFC) voxels that fell only within the resection cavity mask.

### Statistical analyses

Statistical analyses were performed in Rstudio (version 4.3.2).

To test whether the number of resected HFC voxels was related to neurological decline, logistic regressions were performed for both timepoints (SFU and LFU) separately. Another logistic regression was run to test whether the number of resected HFC voxels related to LFU cognitive decline.

Relevant covariates for each of these regressions were identified using Pearson correlation tests (for continuous variables) or T-tests or Mann-Whitney U tests (for categorical variables). If *P* < .1 the covariate was included in the regression model. In case of multicollinearity, the covariate that correlated most strongly with the dependent variable was used. Covariates considered for SFU neurological outcome were molecular subtype, sex, age, preoperative neurological status, preoperative KPS, tumor lateralization, tumor location in terms of resectability index,^23^ and volume of the tumor and combined tumor/cavity mask. Resectability index represents the likelihood of that location to be resected based on a resection probability map and was used as a finegrained measure for tumor localization in eloquent areas^23^. Covariates considered for LFU neurological outcome were sex, age, preoperative neurological status, preoperative KPS score, tumor lateralization, resectability index, volume of the tumor and combined tumor/cavity mask, oncological treatment, and follow-up time (months between surgery and LFU). For cognitive outcome (which was already adjusted for sex, age and education), we considered as covariates: molecular subtype, tumor lateralization, tumor location in terms of resectability index, volume of the tumor and combined tumor/cavity mask, tumor treatment, epilepsy, follow-up time, and baseline cognitive status.

In our preregistration, we planned to enter IDH mutation status as a covariate in all analyses. However, the composition of our study sample was such that only a few patients had IDH-wildtype glioblastomas, prohibiting adequate adjusting for this variable. As such, we deviated from our preregistration and performed separate analyses in the whole group and IDH-mutant glioma patients instead. Our preregistration also envisioned repeating analyses with significant results using stricter inclusion criteria for the timing of LFU, i.e., < 15 months of follow-up time, but the number of patients outside this window was low. Therefore, we opted to not add this sensitivity analysis.

The level of significance of all analyses was set at *P* < 0.05.

## Results

### Participant characteristics

In total, 54 patients met our inclusion criteria (see Table 1). Most patients had IDH-mutant glioma (N = 46, 85%) in the left hemisphere (N = 35, 67%, for tumor occurrence map see Supplementary Figure 2). At BL, 7 patients (13%) had neurological deficits and 18 patients (33%) were cognitively impaired.

**Table 1.** Participant characteristics.

| Characteristic | N = 54 |
| --- | --- |
| **Patient characteristics** |  |
| Sex, male / female | 37 / 17 |
| Age at baseline, years, mean ± SD | 42.1 ± 12.4 |
| Education level (Verhage 2/3/4/5/6/7) | 5 / 3 / 7 / 13 / 13 / 13 |
| Epilepsy at baseline, yes / no | 50 / 4 |
| AED use, yes / no | 49 / 5 |
| Interval resection–NPA, months, mean ± SD | 12.6 ± 3.7 |
| Interval resection–long-term neurological outcome, months, mean ± SD | 13.6 ± 4.5 |
| KPS at baseline, median (IQR) | 100 (90–100) |
| **Treatment before LFU** |  |
| Awake surgery, yes / no / missing | 40 / 13 / 1 |
| Radiotherapy only | 6 |
| Chemotherapy and radiotherapy | 18 |
| None | 30 |
| **Progression before LFU** |  |
| Cognitive outcome, yes / no | 6 / 48 |
| Neurological outcome, yes / no / missing<sup>a</sup> | 7 / 45 / 2 |
| **Tumor characteristics** |  |
| Lateralization, left / right | 36 / 18 |
| Histopathological diagnosis, n |  |
| Glioblastoma, IDH-wildtype | 8 |
| Astrocytoma, IDH-mutant, 1p/19q non-codeleted | 27 |
| Oligodendroglioma, IDH-mutant, 1p/19q codeleted | 19 |
| Tumor volume, mL, mean ± SD<sup>b</sup> | 48.9 ± 36.3 |

<sup>a</sup> Missing values reflect absence of the neurological assessment at 1-year follow-up.
<sup>b</sup> Tumor volume based on the tumor mask including the T2-hyperintense region.
*Abbreviations:* AED, antiepileptic drug; *IDH*, isocitrate dehydrogenase; IQR, interquartile range; KPS, Karnofsky Performance Status; NPA, neuropsychological assessment; SD, standard deviation.

### Neurological outcome at short-term follow-up

For neurological outcome at SFU, all 54 patients could be included, of which 26 patients (48%) showed neurological decline. Testing of potential covariates did not yield any significant associations (*P* > .1).

Logistic regression did not show a significant difference in neurological outcome at SFU according to the number of resected HFC voxels (odds ratio = 0.975, *P* = 0.499, Figure 2A, Supplementary Table S3). Since we only had few IDH-wildtype glioblastoma patients, we repeated these analyses in the IDH-mutant patients only, which also yielded non-significant results (odds ratio = 0.991, P = 0.656, N = 46, Supplementary Table S4).

**Figure 2.**
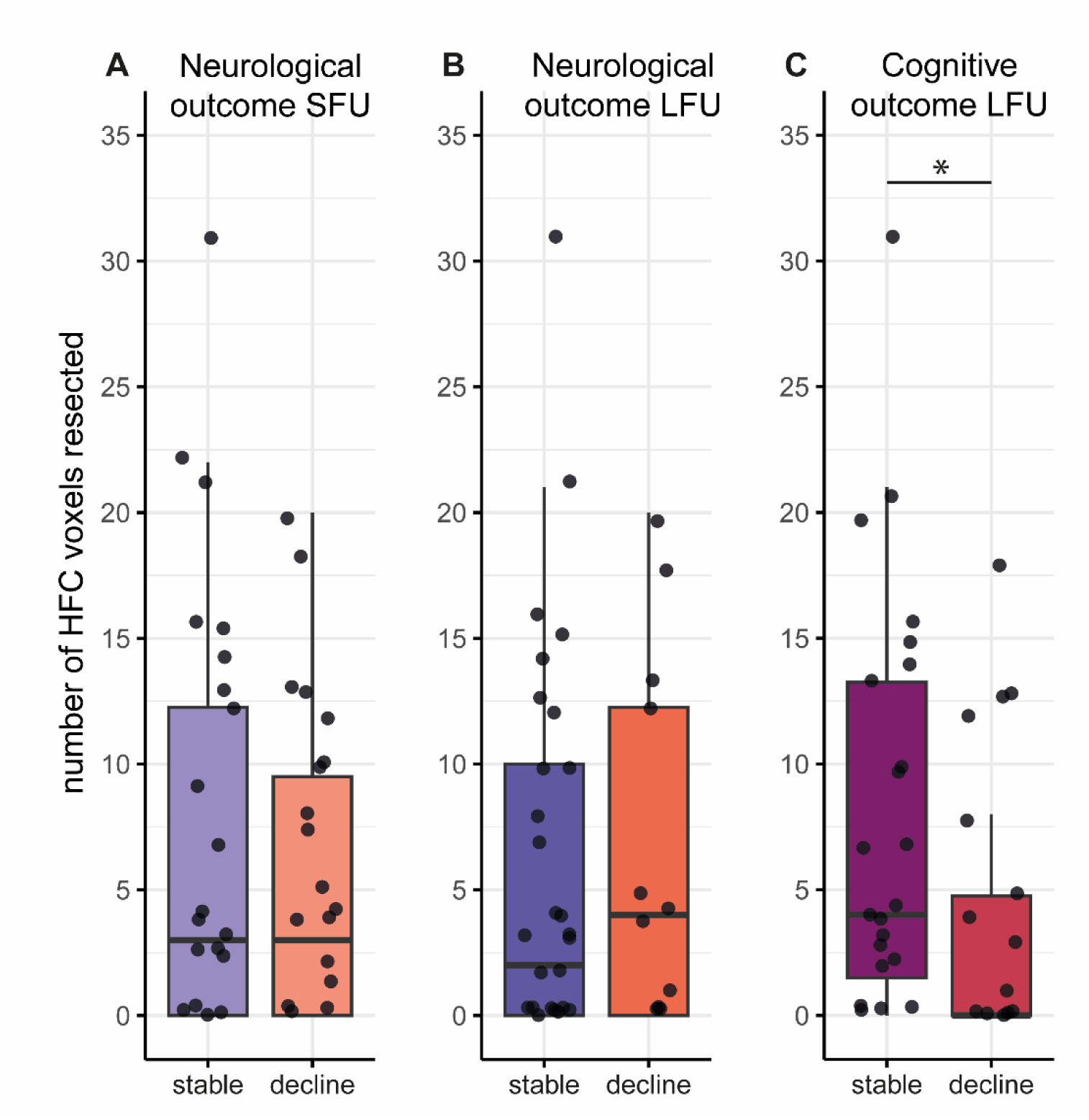
Neurological and cognitive outcomes. The number of resected HFC voxels did not differ between neurological stable or declined status (A) at short-term follow-up (SFU) (B) or long-term follow-up (LFU) (C). Resecting fewer HFC voxels related to cognitive decline at long-term follow-up (C).

We also explored whether the total number of HFC voxels in the merged mask (opposed to solely the resected voxels) related to neurological outcome at SFU, which it did not (odds ratio = 0.994, *P* = 0.644, Supplementary Table S3). Exploratorily, the number of resected voxels with low FC (LFC) also did not relate to the SFU neurological outcome (odds ratio = 0.968, *P* = 0.156). We observed similar patterns of FC for the different neurological scores at SFU (instead of using the delta score categorization into stable versus declining), namely a visual tendency for lower neurological deficits at SFU in the case of a higher number of resected or total voxels with either low or high FC (Supplementary Figure 3). This tendency was statistically non-significant, as was the association between tumor and resection volume and neurological outcome at SFU (Supplementary Figure 4). When applying a logistic regression on the absolute (instead of delta) neurological score at SFU, grouped into no deficits (score = 0) or neurological deficits present (score > 0), we found similar non-significant results (odds ratio = 0.969, *P* = 0.435, Supplementary Table S5).

The distribution of the number of resected HFC voxels was skewed, and there was a significant correlation between the number of HFC voxels resected and tumor volume (Spearman’s rho = 0.53, *P* < 0.001). In post hoc analyses, we therefore also tested the association between a binarized HFC variable (any HFC voxels resected yes or no) and neurological outcome, which would not be impacted by tumor volume. No significant relation was observed between binarized HFC and neurological outcome at SFU (Supplementary Table S3).

### Neurological outcome at long-term follow-up

For neurological outcome at LFU, 45 patients were available (2 had missing scores at LFU, and 7 had tumor progression before LFU), of which 12 patients (27%) declined. Sex and resectability index were found to be relevant covariates and were added to the analyses.

Logistic regression did not show a significant difference in neurological outcome at LFU (stable versus decline) according to the number of resected HFC voxels (odds ratio = 0.991, *P* = 0.683, Figure 2B, Supplementary Table S3). Binarized HFC did not relate either to neurological LFU outcome (odds ratio = 1.158, *P* = 0.861, Supplementary Table S3). Similarly, non-significant findings were present in the IDH-mutant patients only (odds ratio = 0.961, *P* = 0.545, N = 40, Supplementary Table S4).

The total number of resected and non-resected HFC voxels in the merged mask did not relate to neurological outcome at LFU in the full sample (odds ratio = 0.991, *P* = 0.683, Supplementary Table S3), and neither did the number of resected LFC voxels (odds ratio = 1.006, *P* = 0.836). We again observed similar patterns of FC for the different neurological scores at LFU with less neurological deficits in the case of a higher number of resected or total voxels with either low or high FC (Supplementary Figure S3). Again, dichotomizing the absolute neurological outcome score into presence or absence of deficits at LFU resulted in non-significant associations with HFC (odds ratio = 0.998, *P* = 0.971, Supplementary Table S5).

### Cognitive outcome at long-term follow-up

For cognitive outcomes, 46 patients were included (2 had > 6 missing subtest scores at LFU, 5 had tumor progression before LFU, and 1 had both). Of these, 22 patients (48%) showed cognitive decline at LFU. Covariates related to the binary outcome of stable versus declining cognition were preoperative KPS and resectability index.

Logistic regression analyses showed a significant relationship between FC and cognitive decline (odds ratio = 0.854, *P* = 0.023, Figure 2C, Table 1), such that a lower number of resected HFC voxels related to cognitive decline. Two illustrative cases are shown in Figure 3. Results were similar when only testing within the IDH-mutant glioma patients (odds ratio = 0.858, *P* = 0.029, Supplementary Table S6).

**Figure 3.**
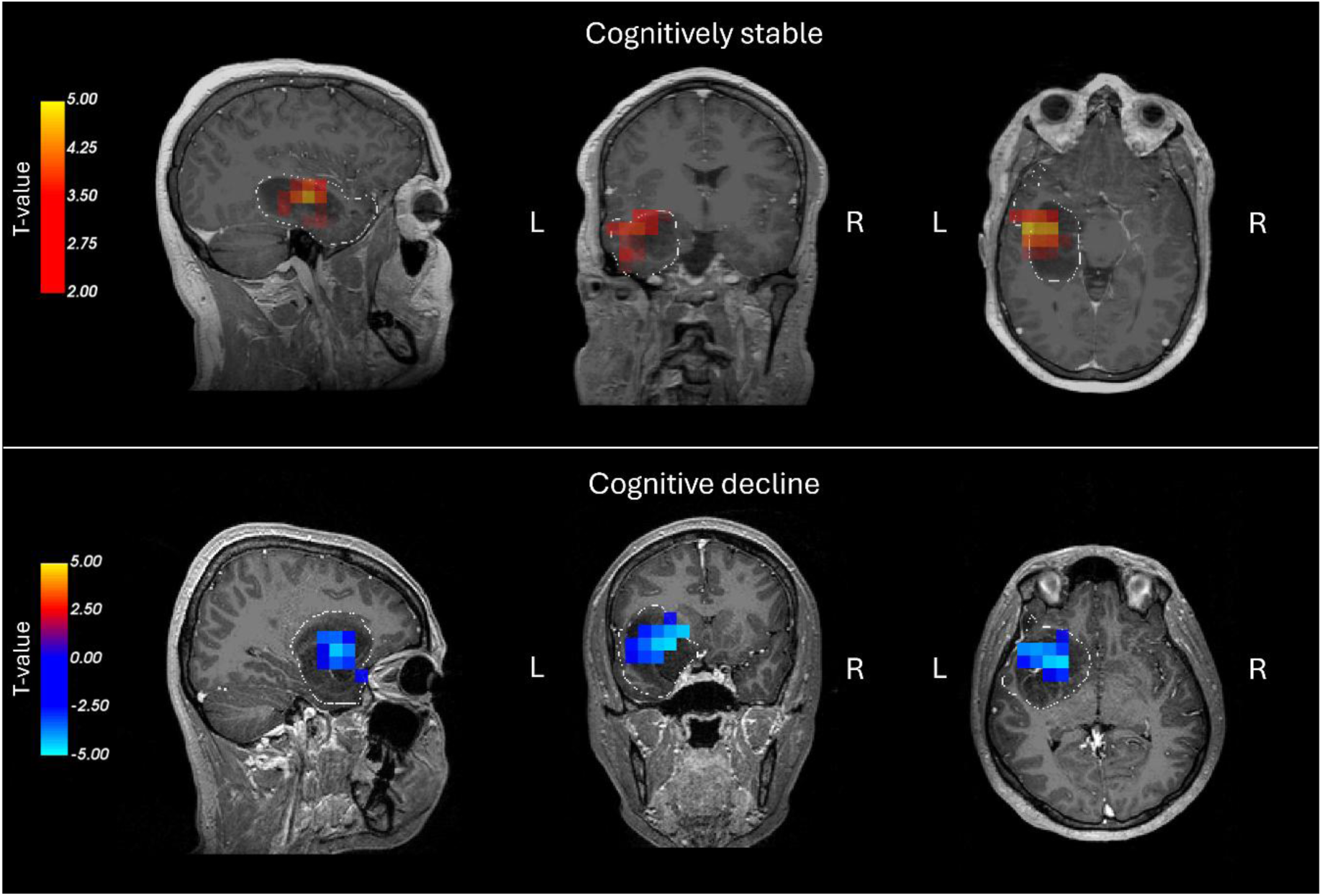
Illustrative cases of preoperative MEG functional connectivity maps in a cognitively stable patient and a patient with cognitive decline. High (red, positive T-values) and low (blue, negative T-values) functional connectivity voxels are superimposed on the preoperative T1-weighted MRI. The white dotted line indicates the tumor margin. Both patients had an astrocytoma (WHO grade II, IDH-mutant, 1p/19q non-codeleted). The cognitively stable patient (tumor volume = 41 mL) had 21 HFC voxels resected, compared to zero resected HFC voxels in the patient with cognitive decline (tumor volume = 55 mL).

Testing the binarized HFC variable (HFC voxels resected yes or no) and cognitive outcome, yielded similar results to the number of HFC voxels resected (odds ratio = 0.076, *P* = 0.011, Table 2). These results indicate that tumor volume did not drive the association between the number of resected HFC voxels and cognition.

**Table 2.**
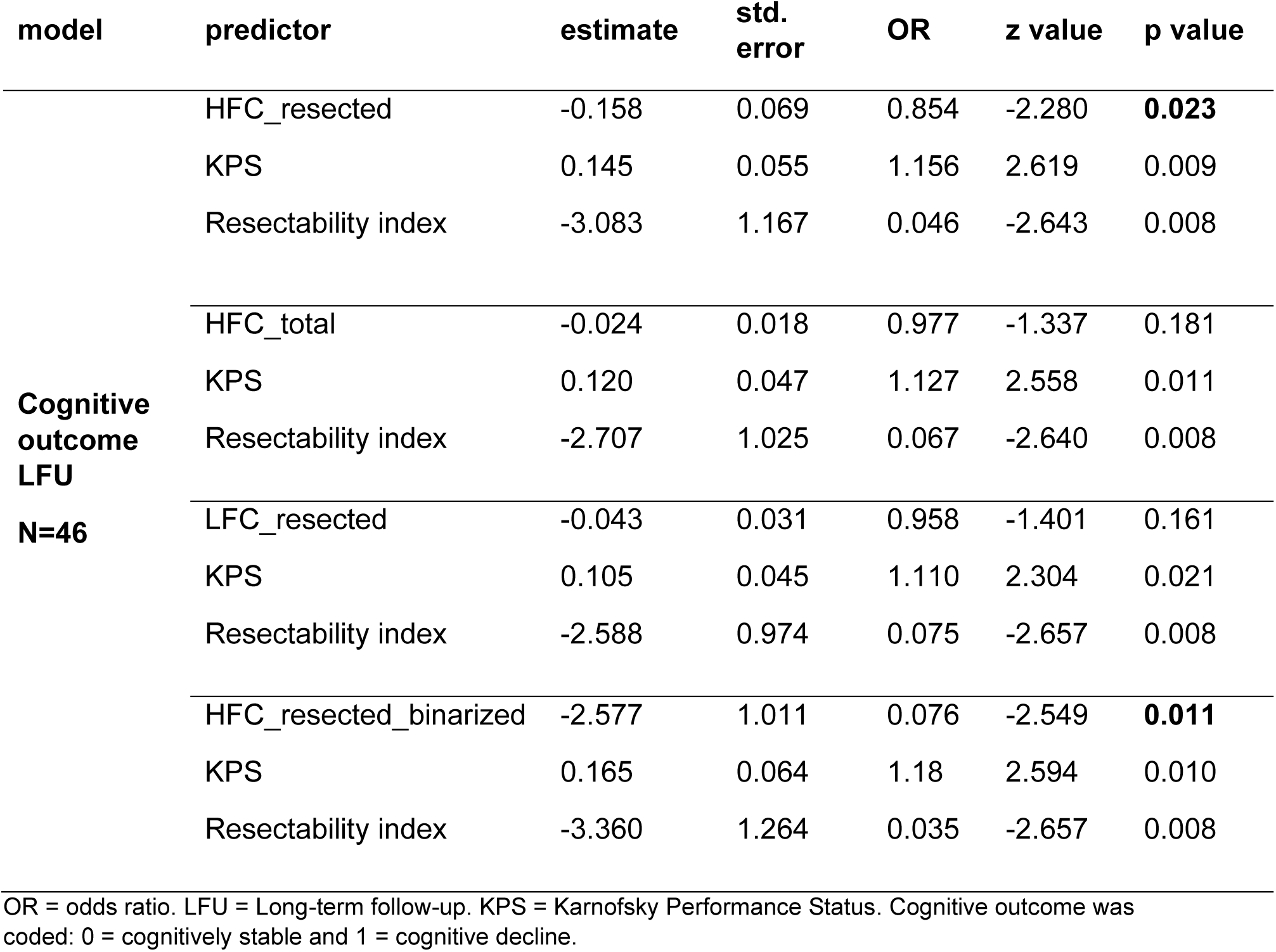
Cognitive outcome regression results.

| model | predictor | estimate | std.<br>error | OR | z value | p value |
| --- | --- | --- | --- | --- | --- | --- |
| **Cognitive**<br>**outcome**<br>**LFU**<br>**N=46** | HFC_resected | -0.158 | 0.069 | 0.854 | -2.280 | **0.023** |
|  | KPS | 0.145 | 0.055 | 1.156 | 2.619 | 0.009 |
|  | Resectability index | -3.083 | 1.167 | 0.046 | -2.643 | 0.008 |
|  | HFC_total | -0.024 | 0.018 | 0.977 | -1.337 | 0.181 |
|  | KPS | 0.120 | 0.047 | 1.127 | 2.558 | 0.011 |
|  | Resectability index | -2.707 | 1.025 | 0.067 | -2.640 | 0.008 |
|  | LFC_resected | -0.043 | 0.031 | 0.958 | -1.401 | 0.161 |
|  | KPS | 0.105 | 0.045 | 1.110 | 2.304 | 0.021 |
|  | Resectability index | -2.588 | 0.974 | 0.075 | -2.657 | 0.008 |
|  | HFC_resected_binarized | -2.577 | 1.011 | 0.076 | -2.549 | **0.011** |
|  | KPS | 0.165 | 0.064 | 1.18 | 2.594 | 0.010 |
|  | Resectability index | -3.360 | 1.264 | 0.035 | -2.657 | 0.008 |

OR = odds ratio. LFU = Long-term follow-up. KPS = Karnofsky Performance Status. Cognitive outcome was coded: 0 = cognitively stable and 1 = cognitive decline.

Exploratorily, the number of LFC resected voxels did not relate to cognitive outcome (odds ratio = 0.958, *P* = 0.161). Finally, we also explored whether the total number of HFC voxels in the combined, merged mask was related to cognitive outcome. This proved not to be the case (odds ratio = 0.977, *P* = 0.181).

## Discussion

We set out to explore the relevance of preoperative, non-invasive MEG-based FC mapping for long-term postoperative cognitive and neurological outcomes. Contrary to our hypothesis, resecting more highly functionally connected (HFC) tumor voxels was associated with long-term cognitive stability: patients with more resected HFC voxels were more often cognitively preserved one year after surgical resection, than those with fewer or no resected HFC voxels. No associations were present between HFC voxel resection and neurological outcomes.

We found that resecting any or more HFC voxels relates to more likely cognitive preservation one year after glioma resection, as compared to not resecting any or resecting fewer HFC voxels. Studies from a different MEG center previously showed a consistent picture regarding the presence of HFC voxels around the surgical timepoint: these are usually functionally relevant and correspond to DES-positive locations in gliomas of all grades^3–5^. Moreover, these voxels contained the most malignant tumor cells, specifically in IDH-wildtype glioblastoma patients.^2^ In terms of cognitive relevance, earlier work found that the removal of HFC voxels resulted in early postoperative decline in language functioning.^7^ However, this decline proved transient, with all HFC resection-related language deficits resolving within three months after surgical resection. This renders our results all the more interesting: resection of HFC voxels may relate to transiently impaired cognition in the short-term postoperative period but to better cognitive outcomes in the long term. Speculatively, such long-term cognitive benefits could in part result from greater oncological control, as HFC voxels are putatively more malignant and associated with shorter survival.^2^ Moreover, FC mapping based on resting-state functional MRI has shown that greater connectivity of the tumor with the rest of the brain may impact cognitively relevant resting-state networks.^24^ This is interesting, as the relationship between MEG-based local activity around the tumor, global connectome topology and cognitive functioning is complex.^25,26^ Future work should establish whether resecting more HFC voxels relates to more beneficial long-term postoperative MEG network changes than in patients with fewer or no resected HFC voxels and cognitive decline. The fact that our current results remained significant within the subsample of IDH-mutant glioma patients, for whom long-term cognitive outcome is even more important in light of their better prognosis compared to IDH-wildtype glioblastoma patients, further supports the exploration of preoperative MEG mapping of HFC voxels in this population of patients. Finally, we did observe a counterintuitive effect of higher KPS relating to cognitive decline, but we must underline that all of our patients had a KPS of 70 or higher.

An open question remains how DES mapping results relate to the relationship between HFC voxel resection and long-term cognitive outcome. We did not have sufficient data on DES-negative locations to explore which HFC voxels were functionally eloquent according to perioperative stimulation, which is a shame as the previous study focusing on language outcome found that the HFC voxels with DES-negative mapping were all the more associated with short-term outcome.^7^ We know that the current sample of included patients had relatively high KPS, most often underwent awake DES mapping, and thus were monitored extensively in terms of surgical resection boundaries. It would be interesting to check how often HFC voxels coincided with positive DES mapping, in relation to both short-term and long-term cognitive outcomes. In such cases, using preoperative HFC maps could provide additional information on how to achieve the best onco-functional balance.

In the current work, we found no relationship between HFC voxel resection and neurological outcome at SFU or LFU. This finding again contrasts with a previous study from UCSF where approximately one week after resection, 60% of the patients with HFC voxel resections had new neurological deficits, compared to 29% of patients with LFC voxel resections. In our (Amsterdam) cohort, these rates were 48% for HFC voxel resections and 45% for LFC resections. In the longer term, patients undergoing HFC voxel resections in the UCSF study had a 25% rate of new neurological deficits 6 months after resection, versus 0% for LFC voxel-only resections.^6^ In comparison, our current analysis shows that patients with HFC voxel resections had a 31% rate of neurological deficits one year after resection, versus 21% for LFC voxels-only resections. Thus, while HFC-related rates of neurological decline seem comparable across studies, LFC-related rates seem inflated in our cohort. Several explanations could be applicable. For one, Tarapore and colleagues reported LFC resections in only 18% of patients, compared to 41% in our study. Such differences in the number of lower FC voxels could be due to various aspects: even though patient cohorts in terms of demographics and tumor characteristics seem similar, the study by Tarapore and colleagues included 31% recurrent glioma patients undergoing re-resection, while we only included *de novo* patients undergoing their first resection. We also used different MEG systems with different sensor types. As it stands, our findings suggest that HFC resection indices may be more replicably related to outcomes across different MEG sites and disease phases, compared to LFC.

Several additional limitations should be considered when interpreting our findings. First, this was a single-center study with a modest sample: although 54 patients met our inclusion criteria, the long-term analyses involved a smaller sub-set. Relatedly, our cohort included only 8 patients with IDH-wildtype glioblastoma, which precluded adequate adjustment for molecular subtype and prompted our preregistered-deviating decision to analyze the whole group and the IDH-mutant subgroup separately. Our findings require confirmation in glioblastoma-enriched cohorts before they can be generalized to these patients as well. Second, the observational design precludes causal inference. The association between resection of HFC voxels and preserved long-term cognition does not establish that removing these voxels is in itself protective, and residual confounding remains possible despite covariate adjustment. In particular, the surgical approach as well as postoperative oncological treatment were based on patient and tumor characteristics, which may confound HFC-cognition links. For instance, the number of resected HFC voxels was positively associated with tumor volume. We addressed this by repeating the analysis with a binarized HFC measure, which yielded consistent results and argues against tumor volume as the sole driver, but we cannot exclude confounding by indication or by unmeasured tumor-and patient-level factors. Third, several analyses were exploratory (including those on tumor-mask HFC, LFC voxels, and the continuous neurological scores) and were not corrected for multiple comparisons, so the primary finding on the relationship between HFC voxel resection and long-term cognitive performance warrants independent replication. Finally, some measurements were constrained by the pragmatic, real-world nature of the data. Neurological status was scored retrospectively from medical records rather than through prospective standardized examination at each timepoint; both neurological and cognitive outcomes were additionally reduced to binary decline categories, discarding information contained in the continuous scores.

In conclusion, we find that long-term cognitive preservation relates to more extensive resection of HFC voxels. Prospective, multicenter studies with harmonized acquisition, integrated DES mapping, and adequate glioblastoma representation will be needed to further establish the clinical utility of preoperative resting-state MEG connectivity mapping.

## Data Availability

Data can be made available in processed and anonymized form under reasonable request.

## Supplementary Material

Is available, see other document.

## Author Contributions

Conceptualization and experimental design: M.R.vL., A.J., S.S.N, V.J., M.K., N.V., P.C.deW.H, A.H., L.D.; Analysis pipeline construction: M.R.vL., A.J., S.S.N, V.J., A.H., L.D.; Software development: A.J.; data collection: M.R.vL., M.K., N.V., P.C.deW.H, A.H., L.D.. All authors reviewed and approved the manuscript.

## References

1. De Witt Hamer PC, Robles SG, Zwinderman AH, Duffau H, Berger MS. Impact of intraoperative stimulation brain mapping on glioma surgery outcome: a meta-analysis. J Clin Oncol. 2012; 30(20):2559–2565.

2. Krishna S, Choudhury A, Keough MB, et al. Glioblastoma remodelling of human neural circuits decreases survival. Nature. 2023; 617(7961):599–607.

3. Tarapore PE, Tate MC, Findlay AM, et al. Preoperative multimodal motor mapping: a comparison of magnetoencephalography imaging, navigated transcranial magnetic stimulation, and direct cortical stimulation. J Neurosurg. 2012; 117(2):354–362.

4. Martino J, Honma SM, Findlay AM, et al. Resting functional connectivity in patients with brain tumors in eloquent areas. Ann Neurol. 2011; 69(3):521–532.

5. Guggisberg AG, Honma SM, Findlay AM, et al. Mapping functional connectivity in patients with brain lesions. Ann Neurol. 2008; 63(2):193–203.

6. Tarapore PE, Martino J, Guggisberg AG, et al. Magnetoencephalographic imaging of resting-state functional connectivity predicts postsurgical neurological outcome in brain gliomas. Neurosurgery. 2012; 71(5):1012–1022.

7. Lee AT, Faltermeier C, Morshed RA, et al. The impact of high functional connectivity network hub resection on language task performance in adult low- and high-grade glioma. J Neurosurg. 2020; 134(3):1102–1112.

8. Louis DN, Perry A, Wesseling P, et al. The 2021 WHO Classification of Tumors of the Central Nervous System: a summary. Neuro Oncol. 2021; 23(8):1231–1251.

9. Karnofsky DA, Abelmann WH, Craver LF, Burchenal JH. The use of the nitrogen mustards in the palliative treatment of carcinoma. Cancer 1948; 1(4):634–656.

10. Lyden P, Brott T, Tilley B, et al. Improved reliability of the NIH Stroke Scale using video training. NINDS TPA Stroke Study Group. Stroke. 1994; 25(11):2220–2226.

11. Douw L, Klein M, Fagel SS, et al. Cognitive and radiological effects of radiotherapy in patients with low-grade glioma: long-term follow-up. Lancet Neurol. 2009; 8(9):810–818.

12. van Lingen MR, Breedt LC, Geurts JJG, et al. The longitudinal relation between executive functioning and multilayer network topology in glioma patients. Brain Imaging Behav. 2023; 17(4):425–435.

13. Gorter M, Rottgering JG, Belgers V, et al. Distinct and reproducible neurocognitive profiles in stable diffuse glioma: A data-driven approach to understanding cognitive heterogeneity. Neuro Oncol. 2025; 27(12):3250–3259.

14. Wefel JS, Vardy J, Ahles T, Schagen SB. International Cognition and Cancer Task Force recommendations to harmonise studies of cognitive function in patients with cancer. Lancet Oncol. 2011; 12(7):703–708.

15. Pemberton HG, Wu J, Kommers I, et al. Multi-class glioma segmentation on real-world data with missing MRI sequences: comparison of three deep learning algorithms. Scientific Reports. 2023; 13(1).

16. Helland RH, Ferles A, Pedersen A, et al. Segmentation of glioblastomas in early post-operative multi-modal MRI with deep neural networks. Sci Rep. 2023; 13(1):18897.

17. Taulu S, Simola J, Kajola M. Applications of the signal space separation method. IEEE Transactions on Signal Processing. 2005; 53(9):3359–3372.

18. Gramfort A, Luessi M, Larson E, et al. MNE software for processing MEG and EEG data. Neuroimage. 2014; 86:446–460.

19. Fischl B. FreeSurfer. Neuroimage. 2012; 62(2):774–781.

20. Hamalainen MS, Sarvas J. Realistic conductivity geometry model of the human head for interpretation of neuromagnetic data. IEEE Trans Biomed Eng. 1989; 36(2):165–171.

21. Sekihara K, Hild KE, Dalal SS, Nagarajan SS. Performance of prewhitening beamforming in MEG dual experimental conditions. IEEE Trans Biomed Eng. 2008; 55(3):1112–1121.

22. Nolte G, Bai O, Wheaton L, Mari Z, Vorbach S, Hallett M. Identifying true brain interaction from EEG data using the imaginary part of coherency. Clin Neurophysiol. 2004; 115(10):2292–2307.

23. Hendriks EJ, Idema S, Hervey-Jumper SL, et al. Preoperative Resectability Estimates of Nonenhancing Glioma by Neurosurgeons and a Resection Probability Map. Neurosurgery. 2019; 85(2):E304–E313.

24. Mandal AS, Wiener C, Assem M, et al. Tumour-infiltrated cortex participates in large-scale cognitive circuits. Cortex. 2024; 173:1–15.

25. Zimmermann MLM, Breedt LC, Centeno EGZ, et al. The relationship between pathological brain activity and functional network connectivity in glioma patients. J Neurooncol. 2024; 166(3):523–533.

26. Zimmermann MLM, Ulrich C, Breedt LC, et al. The relationship between deviant brain activity and executive functioning in glioma patients. Neurooncol Adv. 2025; 7(1):vdaf168.

