## Supplemental_Material for "Resection of highly functionally connected glioma regions predicts long-term cognitive preservation"

### Supplementary Materials

**Table S1** Overview of earlier studies using an overlapping patient cohort

| Author (Year) | Title |
|---|---|
| **Douw et al. (2010) [1]** | Epilepsy is related to theta band brain connectivity and network topology in brain tumor patients |
| **van Dellen et al. (2012) [2]** | MEG network differences between low- and high-grade glioma related to epilepsy and cognition |
| **Van Dellen et al. (2012) [3]** | Connectivity in MEG resting-state networks increases after resective surgery for low-grade glioma and correlates with improved cognitive performance |
| **van Dellen et al. (2014) [4]** | Epilepsy surgery outcome and functional network alterations in longitudinal MEG: a minimum spanning tree analysis |
| **Carbo et al. (2017) [5]** | Dynamic hub load predicts cognitive decline after resective neurosurgery |
| **Derks et al. (2018) [6]** | Oscillatory brain activity associates with neuroligin-3 expression and predicts progression free survival in patients with diffuse glioma |
| **Derks et al. (2019) [7]** | Understanding cognitive functioning in glioma patients: The relevance of IDH-mutation status and functional connectivity |
| **Belgers et al. (2020) [8]** | Postoperative oscillatory brain activity as an add-on prognostic marker in diffuse glioma |
| **Numan et al. (2021) [9]** | Non-invasively measured brain activity and radiological progression in diffuse glioma |
| **Derks et al. (2021) [10]** | Understanding global brain network alterations in glioma patients |
| **van Lingen et al. (2023) [11]** | The longitudinal relation between executive functioning and multilayer network topology in glioma patients |
| **Zimmermann et al. (2024) [12]** | The relationship between pathological brain activity and functional network connectivity in glioma patients |
| **Alexandersen et al. (2025) [13]** | Functional connectotomy of a whole-brain model reveals tumor-induced alterations to neuronal dynamics in glioma patients |
| **Zimmermann et al. (2025) [14]** | The relationship between deviant brain activity and executive functioning in glioma patients |
| **Zimmermann et al. (2026) [15]** | Structural network embedding governs peritumor and distant pathological brain activity in glioblastoma |

**Table S2** Overview of neuropsychological tests and their subscores

| **Cognitive test** | **Subscore** |
|---|---|
| Rey Auditory Verbal Learning Test | Total number of words on trial 1–5 |
| | Delayed recall corrected for trial 5 |
| Memory Comparison Test | Intercept |
| | Slope |
| Categoric Word Fluency Test | Total number of animals |
| Stroop Color-Word Test | Card I (*sec*) |
| | Card II (*sec*) |
| | Card III (*sec*) |
| | Card III corrected for Card II (*sec*) |
| Concept Shifting Test | CST A (*sec*) |
| | CST B (*sec*) |
| | CST C shift (*sec*) |
| Letter Digit Substitution Test | LDST oral (*number of items*) |

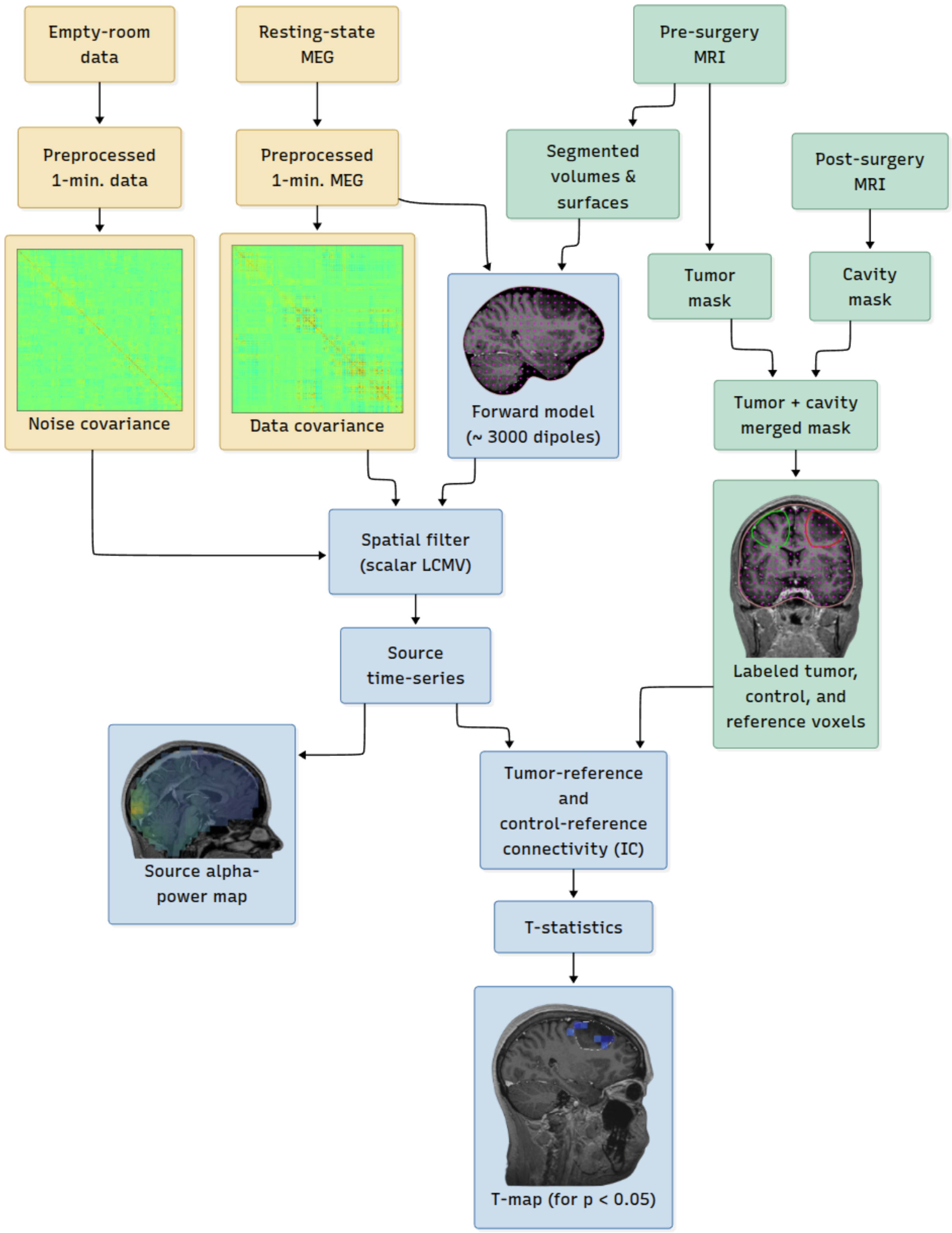


**Supplementary Figure S1.** MEG2map workflow employed in the study.

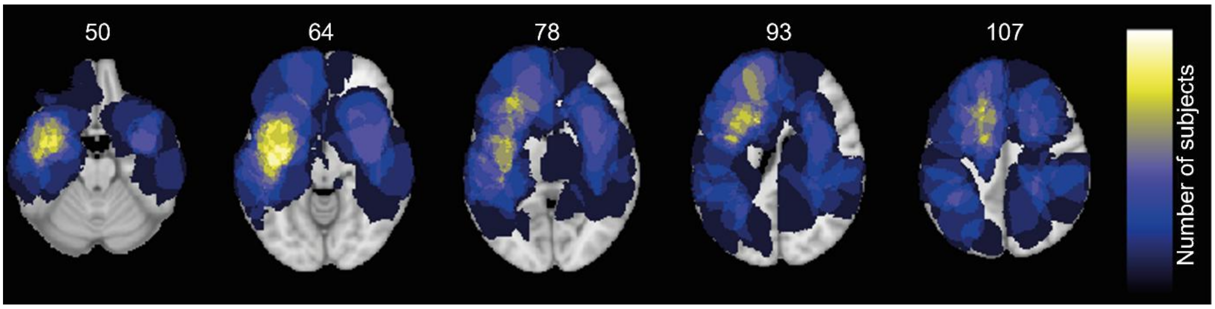


**Supplementary Figure S2.** Lesion map of tumor occurrence.

Colour bar indicates the number of subjects with a tumor in that location. Tumors were more often lateralized in the left hemisphere. Z-value is indicated.

**Table S3** Neurological outcome regression results

| model | predictor | estimate | std. error | OR | z value | p value |
|---|---|---|---|---|---|---|
| **Neurological outcome SFU** **N=54** | HFC_resected | -0.026 | 0.038 | 0.975 | -0.676 | 0.499 |
| | HFC_total | -0.006 | 0.013 | 0.994 | -0.462 | 0.644 |
| | LFC_resected | -0.032 | 0.023 | 0.968 | -1.420 | 0.156 |
| | HFC_resected_binarized | 0.022 | 0.551 | 1.023 | 0.041 | 0.967 |
| **Neurological outcome LFU** **N=45** | HFC_resected | -0.012 | 0.057 | 0.988 | -0.218 | 0.827 |
| | Resectability index | -3.238 | 1.323 | 0.039 | -2.447 | 0.014 |
| | Sex | 2.615 | 1.046 | 13.66 | 2.499 | 0.013 |
| | HFC_total | -0.009 | 0.021 | 0.991 | -0.408 | 0.683 |
| | Resectability index | -3.247 | 1.323 | 0.039 | -2.454 | 0.014 |
| | Sex | 2.612 | 1.017 | 13.63 | 2.568 | 0.010 |
| | LFC_resected | 0.006 | 0.031 | 1.006 | 0.207 | 0.836 |
| | Resectability index | -3.236 | 1.317 | 0.039 | -2.458 | 0.014 |
| | Sex | 2.599 | 1.030 | 13.45 | 2.523 | 0.012 |
| | HFC_resected_binarized | 0.147 | 0.841 | 1.158 | 0.175 | 0.861 |
| | Resectability index | -3.222 | 1.317 | 0.040 | -2.447 | 0.014 |
| | Sex | 2.508 | 1.026 | 12.28 | 2.443 | 0.015 |

OR = odds ratio. Neurological outcome was coded: 0 = stable and 1 = decline.

**Table S4**. Neurological outcome regression results in IDH mutant only

| model | predictor | estimate | std. error | OR | z value | p value |
|---|---|---|---|---|---|---|
| **Neurological outcome SFU**<br><br>**N=46** | HFC_resected | -0.023 | 0.041 | 0.977 | -0.555 | 0.579 |
| | HFC_total | -0.009 | 0.020 | 0.991 | -0.446 | 0.656 |
| | LFC_resected | -0.046 | 0.026 | 0.954 | -1.829 | 0.068 |
| **Neurological outcome LFU**<br><br>**N=40** | HFC_resected | -0.040 | 0.066 | 0.961 | -0.605 | 0.545 |
| | Resectability index | -3.275 | 1.362 | 0.038 | -2.405 | 0.016 |
| | Sex | 2.731 | 1.129 | 15.347 | 2.419 | 0.016 |
| | HFC_total | -0.005 | 0.028 | 0.995 | -0.184 | 0.854 |
| | Resectability index | -3.200 | 1.324 | 0.041 | -2.417 | 0.016 |
| | Sex | 2.533 | 1.061 | 12.589 | 2.386 | 0.017 |
| | LFC_resected | 0.004 | 0.032 | 1.004 | 0.139 | 0.890 |
| | Resectability index | -3.196 | 1.320 | 0.041 | -2.421 | 0.015 |
| | Sex | 2.522 | 1.067 | 12.458 | 2.365 | 0.018 |

OR = odds ratio. Neurological outcome was coded: 0 = stable and 1 = decline.

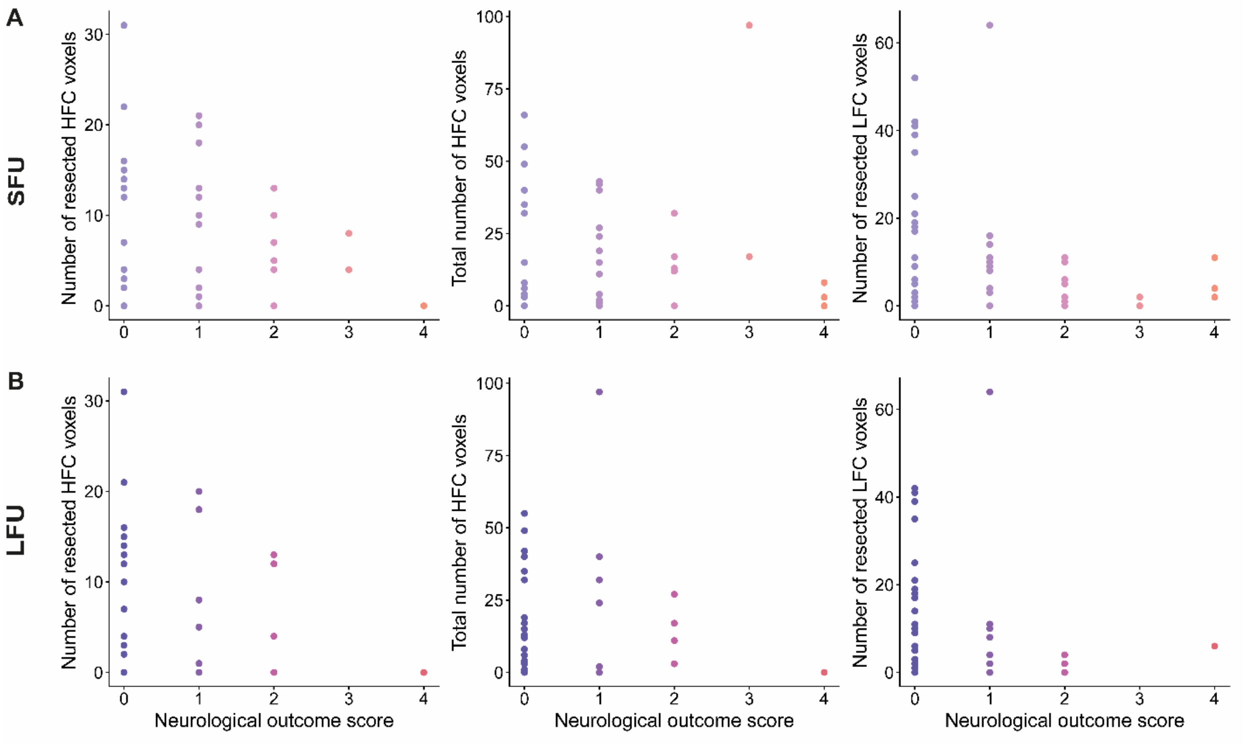


**Supplementary Figure S3.** Absolute neurological outcome score at short term (SFU) and long term follow-up (LFU).

Neurological outcome is the total NIH-SS score. Visual inspection suggests an association between higher numbers of total and resected HFC/LFC voxels and fewer neurological deficits.

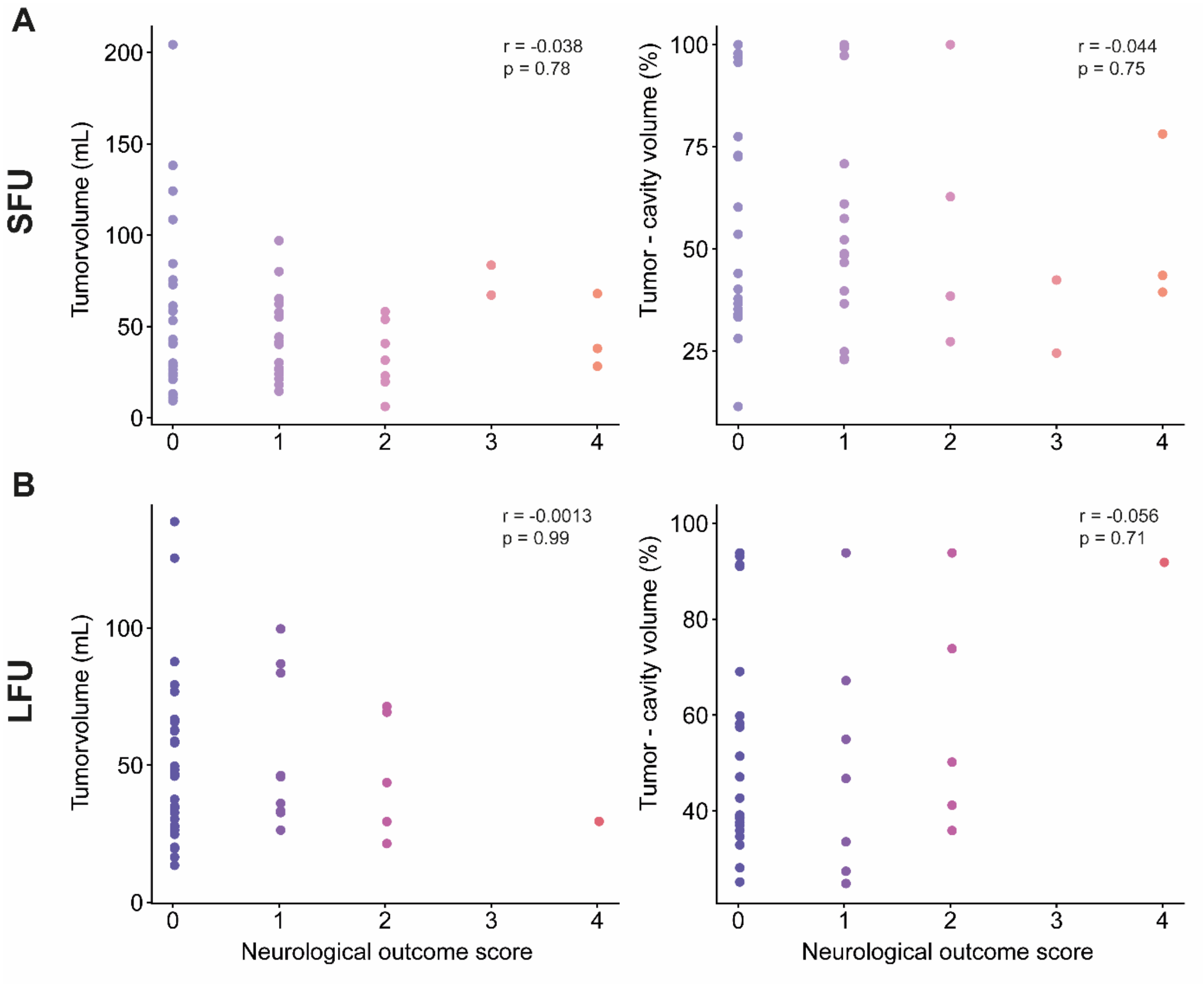


**Supplementary Figure S4.** Tumor volume and extent of resection do not correlate with absolute neurological outcome score at short-term (SFU) and long-term follow-up (LFU). Extent of resection was calculated as (tumor volume – cavity volume) * 100%. Neurological outcome is the total NIH-SS score. A Spearman correlation was run, as the neurological outcome score was not normally distributed.

**Table S5.** Neurological outcome regression results with absolute NIH score categorized into no neurological deficits (NIH=0) or neurological deficits (NIH>0)

| model | predictor | estimate | std. error | OR | z value | p value |
|---|---|---|---|---|---|---|
| **Neurological outcome SFU N=54** | HFC_resected | -0.032 | 0.041 | 0.969 | -0.781 | 0.435 |
| | Sex | 1.470 | 0.692 | 4.349 | 2.125 | 0.034 |
| **Neurological outcome LFU N=45** | HFC_resected | 0.015 | 0.053 | 1.015 | 0.285 | 0.776 |
| | Age | 0.063 | 0.032 | 1.065 | 1.950 | 0.051 |
| | Resectability index | -2.727 | 1.112 | 0.065 | -2.452 | 0.014 |

OR = odds ratio. Neurological outcome was coded: 0 = no deficits and 1 = neurological deficits present.

**Table S6.** Cognitive outcome regression results in IDH mutant only

| model | predictor | estimate | std. error | OR | z value | p value |
|---|---|---|---|---|---|---|
| **Cognitive outcome LFU** | HFC_resected | -0.153 | 0.070 | 0.858 | -2.182 | **0.029** |
| | KPS | 0.127 | 0.054 | 1.135 | 2.332 | 0.020 |
| **N=41** | Resectability index | -2.643 | 1.165 | 0.071 | -2.269 | 0.023 |
| | HFC_total | -0.051 | 0.029 | 0.950 | -1.760 | 0.078 |
| | KPS | 0.111 | 0.048 | 1.117 | 2.303 | 0.021 |
| | Resectability index | -2.433 | 1.078 | 0.088 | -2.256 | 0.024 |
| | LFC_resected | -0.044 | 0.031 | 0.957 | -1.401 | 0.161 |
| | KPS | 0.089 | 0.045 | 1.094 | 1.994 | 0.046 |
| | Resectability index | -2.296 | 0.986 | 0.101 | -2.328 | 0.020 |

OR = odds ratio. Cognitive outcome was coded: 0 = cognitively stable and 1 = cognitive decline.

#### References


1. Douw, L., et al., *Cognitive and radiological effects of radiotherapy in patients with low-grade glioma: long-term follow-up.* Lancet Neurol, 2009. **8**(9): p. 810–8.
2. van Dellen, E., et al., *MEG network differences between low- and high-grade glioma related to epilepsy and cognition.* PLoS One, 2012. **7**(11): p. e50122.
3. van Dellen, E., et al., *Connectivity in MEG resting-state networks increases after resective surgery for low-grade glioma and correlates with improved cognitive performance.* Neuroimage Clin, 2012. **2**: p. 1–7.
4. van Dellen, E., et al., *Epilepsy surgery outcome and functional network alterations in longitudinal MEG: a minimum spanning tree analysis.* Neuroimage, 2014. **86**: p. 354–63.
5. Carbo, E.W., et al., *Dynamic hub load predicts cognitive decline after resective neurosurgery.* Sci Rep, 2017. **7**: p. 42117.
6. Derks, J., et al., *Oscillatory brain activity associates with neuroligin-3 expression and predicts progression free survival in patients with diffuse glioma.* J Neurooncol, 2018. **140**(2): p. 403–412.
7. Derks, J., et al., *Understanding cognitive functioning in glioma patients: The relevance of IDH-mutation status and functional connectivity.* Brain Behav, 2019. **9**(4): p. e01204.
8. Belgers, V., et al., *Postoperative oscillatory brain activity as an add-on prognostic marker in diffuse glioma.* J Neurooncol, 2020. **147**(1): p. 49–58.
9. Numan, T., et al., *Non-invasively measured brain activity and radiological progression in diffuse glioma.* Sci Rep, 2021. **11**(1): p. 18990.
10. Derks, J., et al., *Understanding Global Brain Network Alterations in Glioma Patients.* Brain Connect, 2021. **11**(10): p. 865–874.
11. van Lingen, M.R., et al., *The longitudinal relation between executive functioning and multilayer network topology in glioma patients.* Brain Imaging Behav, 2023. **17**(4): p. 425–435.
12. Zimmermann, M.L.M., et al., *The relationship between pathological brain activity and functional network connectivity in glioma patients.* J Neurooncol, 2024. **166**(3): p. 523–533.
13. Alexandersen, C.G., et al., *Functional connectotomy of a whole-brain model reveals tumor-induced alterations to neuronal dynamics in glioma patients.* Netw Neurosci, 2025. **9**(1): p. 280–302.
14. Zimmermann, M.L.M., et al., *The relationship between deviant brain activity and executive functioning in glioma patients.* Neurooncol Adv, 2025. **7**(1): p. vdaf168.
15. Zimmermann, M.L., et al., *Structural network embedding governs peritumor and distant pathological brain activity in glioblastoma.* medRxiv, 2026: p. 2026.05.05.26352433.
